# Protocol for partnering with peers intervention to improve medication adherence among African Americans with Type 2 Diabetes

**DOI:** 10.1101/2020.06.04.20122895

**Authors:** Olayinka O Shiyanbola, Martha Maurer, Earlise C Ward, Lisa Sharp, Jonas Lee, Adati Tarfa

**Author notes:** These authors contributed equally to this work (Olayinka O Shiyanbola, Martha Maurer, Earlise C Ward, Lisa Sharp, Jonas Lee). This author also contributed to this work (Adati Tarfa). Corresponding author: (OS).

## Abstract

African Americans (AAs) with diabetes are more likely to develop diabetes-related complications and have the highest diabetes-related mortality rates, than all other racial/ethnic groups. These health disparities are primarily due to poor medication adherence (defined as not taking medications as prescribed). AAs have substantially lower adherence to diabetes medications than whites, which contributes to higher rates of diabetes-related complications, such as amputations and strokes. There is a critical need to develop diabetes self-management interventions that improve medication adherence, clinical outcomes and in turn reduce morbidity and mortality among AAs with diabetes. Focusing on psychosocial factors such as health beliefs, self-efficacy and patient-provider communication is instrumental to improving AAs medication adherence. To address this need, we developed, the Peers Supporting Health Literacy, Self-Efficacy, Self-Advocacy, and Adherence (Peers LEAD) intervention, which provides AAs with culturally adapted diabetes and medication beliefs information, one-on-one peer support from AAs with diabetes, and communication and self-efficacy skill development to enhance medication adherence. This pilot research is a pre-post single group intervention study design which will be conducted in two phases using a community engaged approach. The objective is to test the Peers LEAD intervention in Phase 1, and then examine specific intervention elements for refinement in Phase 2. We will employ both quantitative and qualitative methods to assess the feasibility, acceptability, and outcomes of Peers LEAD. Building on established community partnerships, we plan to recruit and enroll 30 Peer Buddies and 20 Peer Ambassadors to participate in the intervention. By utilizing patient feedback to refine Peers LEAD and piloting it to examine its feasibility, we will generate evidence regarding its real world use and provide support for a randomized controlled trial of its impact on AAs diabetes medication adherence and clinical outcomes.

## Introduction

One in six African Americans (AAs) are diagnosed with diabetes at a significantly higher rate than non-Hispanic whites.[1] The rate of diabetes-related complications are also high among AAs. For example, the death rate due to diabetes for AAs is 27% higher than among non-Hispanic whites.[2] Also, diabetes-related kidney disease, blindness and amputations occur four times more among AAs compared to other races.[3] While there are no definitive answers to the cause of these diabetes disparities, differences in optimal health behaviors among AAs contributes to it. [4] Diabetes self-management behavior that reduces disease complications remains a challenge for AAs, possibly due to culturally misaligned care. Diabetes complications can be mitigated by adhering to treatment recommendations.[5–7]

Racial and ethnic minorities have poorer treatment adherence compared to whites.[4] Health disparities among minorities could occur possibly because of cultural differences between patient and providers in the perception of illness and medicine, and is a reflection of the disparities in adherence to medical regimens.[8] In one study, AAs with type 2 diabetes had a 12% lower medication adherence rate compared to whites, leading to more complications and disability.[6] As well, AAs had substantially lower adherence to diabetes medications than whites, though the number of physician encounters and A1c tests, and continuity of care did not differ between them.[9] AAs with diabetes may have comparatively poorer outcomes compared to whites because of underlying unique cultural beliefs and practices, low self-efficacy related to taking medicines and low engagement which influence patient-provider interactions.[6, 9]

Focusing on psychosocial factors such as health beliefs, self-efficacy and patient-provider communication is instrumental to improving AAs medication adherence.[10–12] In our prior studies, patients differed in their perceptions of the need for their medication and their concerns regarding the medication’s adverse effects.[13–18] Evidence shows that health providers do intervene on illness and medication beliefs to improve medication adherence[19, 20] and these interventions work.[21, 22] Patient illness and medication beliefs also influence treatment adherence in AAs with diabetes,[23, 24] therefore, interventions to modify these beliefs towards improving diabetes medication adherence among AAs are critical. Prior work suggests that beliefs and self-efficacy are modifiable in interventions for AAs within a short time frame.[25, 26] Also, among AAs with diabetes, social support is critical.[27] Knowing the influence of these factors on medication adherence, we aimed to develop an intervention that incorporates these factors to improve medication adherence for AAs.

Diabetes self-management can be physically and emotionally taxing for patients.[28–30] Prior diabetes management interventions offer patient support through nurse phone calls or community health workers visits; however, these require expensive and resource intensive professional support staff.[31, 32] Peer mentors represent a more informal, practical, flexible and low-cost means of providing one-on-one support with potentially similar benefits.[33] Peer interactions between individuals with diabetes provide continuous informational and emotional support, and mutual reciprocity, which leads to improved diabetes care.[32, 34] Peer supporters link individuals to diabetes care and encourage/motivate them to take action towards appropriate clinical care.[35, 36] Peers use their own personal experiences with the disease to help others learn how to manage their diabetes successfully. As well, peer support is a practical and sustainable approach to assist with diabetes adherence in disadvantaged communities.[36] For AAs for whom clinic visits frequently fail to improve medication adherence, peer interventions offer a 24/7 community of willing and able supporters of people with diabetes.[37],[32, 38] This study is the first to engage AAs with diabetes who are successfully taking their medicines in enhancing medication adherence for other AAs[24, 39]. Peers offer culturally-appropriate psychosocial support for AAs with diabetes[33] who may distrust the healthcare system.[40] Enhancing patient activation skills (stating preferences and concerns, asking questions) can increase AAs engagement in their medication adherence, in ways clinicians cannot through visits alone.[41]

The intervention, Peers Supporting Health Literacy, Self-Efficacy, Self-Advocacy, and Adherence (Peers LEAD) provides AAs with culturally adapted diabetes and medication beliefs information, one-on-one peer support from AAs/Blacks with diabetes, and communication and self-efficacy skill development to enhance medication adherence. In this program, AAs who have been diagnosed with diabetes and are adherent to their medicines (Peer Ambassadors – PAs) are paired with AAs who are also living with diabetes but are nonadherent to their diabetes medicines (Peer Buddies – PBs). The Peers LEAD intervention components include face-to-face group education sessions and phone/mobile app follow-ups of ambassadors with buddies over 8-weeks. The group education will be delivered by a physician, pharmacist, and diabetes educator separately. Ambassadors will address misperceptions about medicines and diabetes, share their experiences managing diabetes and diabetes medicines, as well as actively support and teach about building patient-provider relationships. This paper describes the protocol design and implementation of Peers LEAD, which will be carried out in two cities in a US Midwestern State.

## Materials and Methods

### Research Design

This pilot research is a pre-post single group intervention study design which will be conducted in two phases using a community engaged approach. We will test the Peers LEAD intervention in Phase 1, and then examine specific intervention elements for refinement in Phase 2. We will employ both quantitative and qualitative methods to assess the feasibility, acceptability, and outcomes of Peers LEAD (Fig. 1). The rationale for assessing both outcomes is that neither quantitative nor qualitative methods are sufficient in explaining the outcomes of the intervention. Mixing both methods gives a more complete analysis of Peers LEAD.[42] Qualitative results collected during and after the intervention will allow us to further explain the outcomes, examine participant’s experiences and modify the methods in a follow-up and/or dissemination study. The study is registered at https://clinicaltrials.gov/ct2/show/NCT04028076.

**Figure 1.**
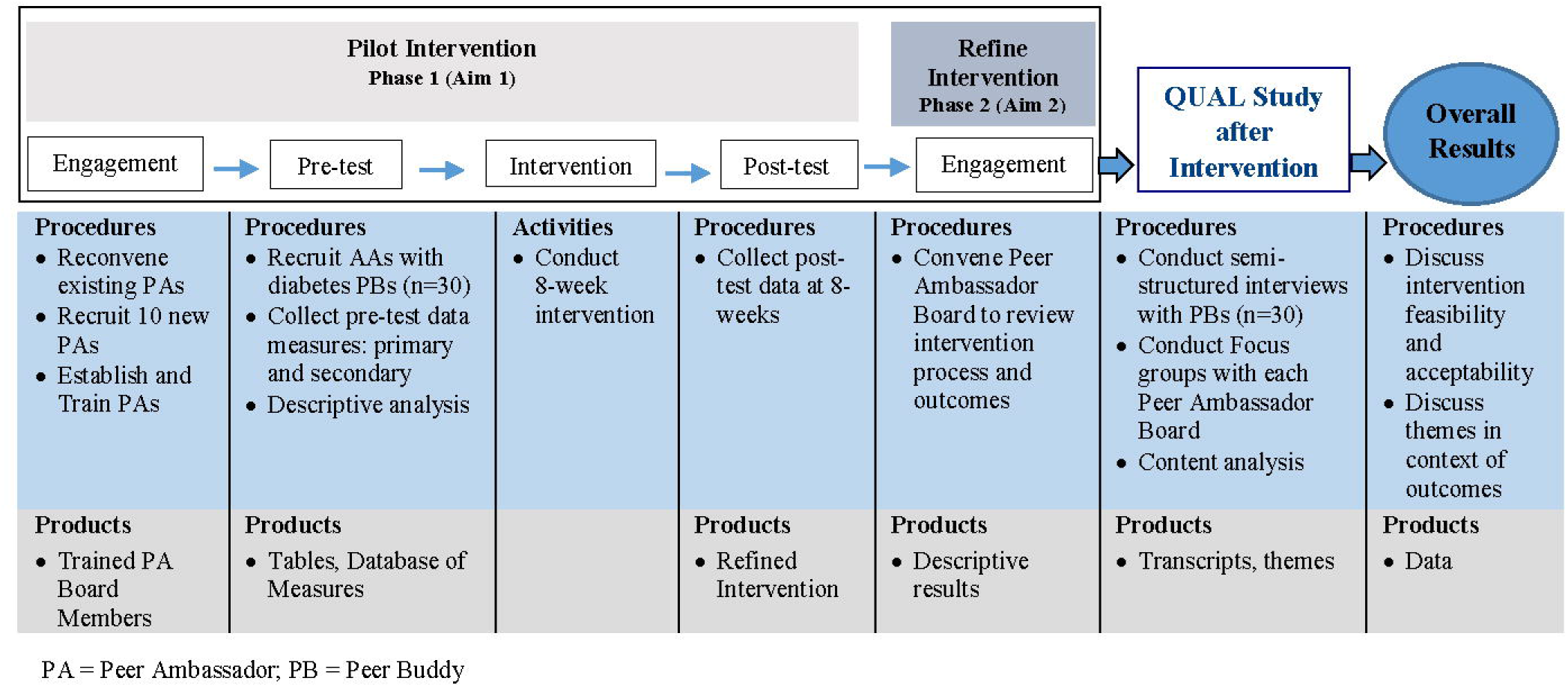
A Community-Engaged Approach to Conduct an Educational Behavioral Intervention to Improve Medication Adherence and Blood Glucose among AAs with Type 2 Diabetes.

### Theoretical and Conceptual Framework

Our prior work utilizing the extended self-regulatory model (ESRM) among AAs with diabetes set the stage for intervention development. The main theoretical frameworks guiding this research are the ESRM and the information-motivation-behavioral skills model (IMB). We chose the IMB as the foundation for this study because it provides a relatively simple, yet powerful explanation for complex health behaviors like medication adherence. Also, it focuses on constructs needed for successful diabetes self-management [43] including *information* “an initial prerequisite for enacting a health behavior”[44], *motivation* that takes into account beliefs about the intervention and attitudes toward adherence[45, 46] and perceived social support for engaging in a behavior,[47] and *behavioral skills* emphasize increasing self-efficacy and activation.[45] The ESRM integrates with the information-belief component of the IMB in this study because it addresses misperceived information on illness and medication beliefs among AAs.[48–51] Peer support fits within a social support model.[52] Social support influences health through psychological and behavioral pathways to improve medication adherence.

### Study Aims and Hypotheses

The study objective is to pilot Peers LEAD to examine for feasibility and outcomes including culturally-informed diabetes-health beliefs, diabetes and medication self-efficacy, patient activation (a measure of engagement/empowerment), medication adherence, and blood glucose, and refine Peers LEAD via participant feedback. The central hypothesis is that Peers LEAD will be feasible and effective for AAs with diabetes, leading to improved outcomes in self-efficacy, activation, adherence, and blood glucose. The study aims are to: (1) pilot Peers LEAD to test the feasibility and acceptability of the intervention intended to shift negative diabetes-health beliefs to positive, enhance diabetes self-efficacy, and improve patient activation, self-reported medication adherence, and blood glucose levels, and (2) refine Peers LEAD for AAs with diabetes using feedback from PAs and PBs.

For Aim 1, a physician, pharmacist and diabetes educator will provide structured education in conjunction with PAs to help deliver Peers LEAD, via initial face-to-face and phone/messaging app follow-ups, serving as PAs. PBs will participate in the program, with a target of 20 PBs in City #1 and 10 PBs in City #2. PBs will complete surveys and hemoglobin (A1c) tests assessing changes in outcomes. Interviews and focus groups will be conducted with all PBs and PAs respectively to get feedback on the intervention.

Aim 2 involves (1) establishing two Peers Ambassador Boards (PABs) one in each of the two cities, (2) training PAB members as PAs, and (3) eliciting feedback to refine Peers LEAD.

### Participants

This study will include two groups of participants: (1) PAs who also serve as our advisory board members and (2) PBs, who are program participants.

#### Peer Ambassadors

PA inclusion criteria are: (1) Men and women 30–65 years old with type 2 diabetes who self-identify as Black/AA and can speak/read English, (2) self-report as prescribed at least one oral diabetes medication and are adherent to these medicines (Score of 11 on the Adherence to Refills and Medications-Diabetes Scale)[53] and (3) have access to and can use a phone with cellular and internet use during the study period.

PA exclusion criteria: Individuals who:(1) are only using injectable insulin; and (2) report diagnosis of serious mental illness/psychosis.

#### Peer Buddies

PBs are program participants who meet the following inclusion criteria: (1) Men and women 30–65 years old with type 2 diabetes who self-identify as Black/AA and can speak and read English. (2) Self-report as prescribed an oral diabetes medication and are nonadherent (Score of greater than 11 on the Adherence to Refills and Medications-Diabetes (ARMS-D) Scale) [53]. (3) Have access to and can use a phone with cellular and internet use during the study period.

Individuals who: (1) are only using insulin, and (2) have a diagnosed psychiatric disorder will be excluded from being a PB.

### Recruitment

Purposive sampling will be used for recruitment of all PAs and PBs.

#### Peer Ambassadors

We will work with our community stakeholders to recruit 10–12 PAs from each site who will constitute the PAB of AAs who are adherent to their medicines and currently successfully managing diabetes. To ensure we can meet our recruitment goal, we will recruit up to 24 individuals, in case some PAs withdraw. We hope to enroll at least 20 PAs (10 in each city).

For one of the cities, where our prior study was conducted[54], recruitment will be accomplished by inviting our established 9-member (3 men, 6 women) PAB to participate in the 8-week intervention. We anticipate that some of the former PAs who participated in the similar pilot 3-week study[54]will likely participate as PAs again, however there will be a need to recruit some additional PAs, so that we have 10 PAB members. For the second city, we will work with our local community stakeholders to recruit 10 PAs who will constitute the second PAB.

#### Peer Buddies

We will work with our community partners in each city to recruit a total of 20 PBs from churches, apartments, and senior centers. Using prior IRB-approved procedures, we will recruit using flyers, personal contacts/word of mouth, and referrals from community partners.

City #1: We will recruit PBs from the following community sites: (1) The University of Wisconsin Collaborative Center for Health Equity connects community partners with University of Wisconsin – Madison faculty working towards improving health equity. In our prior work, they connected the research team to the UW Community Partnership Advisory Board to facilitate the announcement of this recruitment opportunity to community leaders within the city, (2) A local senior center whose mission it is to enhance the ability of seniors to maintain independent lives. Notably, the center has a cultural diversity program for AAs ≥ 55 years old linked to apartment homes and an active AA diabetes support group attended by 20 seniors and their adult caregivers. We will recruit using flyers placed at senior center meeting spaces, as well as by personal contact/word of mouth from their Cultural Diversity Specialist to their diabetes support group, community leaders and affiliated sites including local churches, (3) A faith-based CAB comprised of predominantly AA churches will work with us to publicize this study in the local AA faith community, (4) A local food pantry that has a diabetes wellness program, provides food-insecure patients with free diabetes-appropriate food. They will publicize Peers LEAD through their diabetes program, (5) A study team member who is a UW Department of Family Medicine and Community Health Professor and a Community Health Center Physician will inform potentially eligible patients within his clinical practice about the study opportunity and will then direct the patient to contact the study team if they are interested in learning more.

City #2: Building on past successful collaborations, PBs will be recruited from three different community organizations: (1) a community advisory board (CAB) made up of 15 church leaders, parish nurses, healthcare providers, and social service organization leaders in the city. Peers LEAD was presented to them and they are ready to partner in its implementation. The CAB members will offer ideas/advice on recruitment including the design/content of flyers, potential recruitment locations as well as community contacts that can actively recruit, (2) a church located in a neighborhood with a large population of AAs that has a food pantry that serves 450 people with diabetes. This outreach is coordinated by a nurse with expertise in diabetes management, (3) a federally qualified health center clinic pharmacist will inform potentially eligible patients in her pharmacy clinic about the study opportunity and will then direct the patient to contact the study team if they are interested in learning more. The Health Sciences Institutional Review Board of the primary investigators’ university approved the study.

### Intervention

Peers LEAD is an 8-week culturally adapted peer-supported educational-behavioral intervention that consists of structured group diabetes education and individual phone-based follow-up support with PAs (Table 1). There will be 3 separate group education sessions led by a physician, pharmacist, and diabetes educator. Peer Ambassador Board (PAB) members, who are also PAs will help deliver Peers LEAD, via face-to-face contact with their PBs during group sessions and then phone/messaging app follow-ups. All PAs will attend each group session so that they learn together with their PBs and build social interactions. PAs will be recruited first and then matched to PBs based on gender and age. Weeks 1 and 2 of Peers LEAD consists of 2-hour group education between the participants and diabetes educator, and participants and pharmacist, respectively. Pre-test intervention data will be collected in Week 1. In Weeks 3–7, PAs will deliver the intervention content to participants over the phone as well as help PBs set a goal towards getting healthy. A video/text messaging app, called WhatsApp® will be used to further enhance PA-PB interaction. WhatsApp® is a free cross-platform messaging and Voice over IP app that allows the prompt sharing of text messages, voice calls, video calls, and other media and documents. PAs will use the app to video call PBs 15–30 minutes weekly and actively support and engage with PBs regarding diabetes self-care. Week 8 will focus on a discussion of clinical outcomes by a physician, delivered face-to-face in a concluding group session for all PAs and PBs. Post-test intervention data will also be collected.

**Table 1.** Details of 8-Week Peers LEAD Intervention.

| **Week** | **Format** | **Description** |
| --- | --- | --- |
| **1** | Group education | A diabetes educator targets negative illness beliefs with a focus on the cause and consequence of diabetes. |
| **2** | Group education | A pharmacist reframes medication beliefs to decrease medication concerns and increase necessity in medicines. |
| **3** | Phone | Discuss self-efficacy, positive living and coping with diabetes. |
| **4** | Phone | Provide support for addressing fear, frustration, and emotional distress. |
| **5** | Phone | Discuss self-advocacy in provider communication/relationship building. |
| **6** | Phone | Discuss family/community bonding and maintaining cultural experiences. |
| **7** | Phone | Set a goal for improving health. |
| **8** | Group education | A physician discusses hemoglobin A1C and blood glucose. |

### Training of Peer Ambassadors

To prepare to implement Peers LEAD, PAs will attend a series of 3 meetings, co-facilitated by Wisconsin Network for Research Support (WINRS) and the research team. The first meeting will be an orientation to their role and the second and third meetings will focus on preparing the PAs to implement specific elements of Peers LEAD, such as the phone calls. WINRS will consult with our research team on all PAB meetings. For > 8 years, WINRS staff have been deeply involved in stakeholder engagement, as well as trained, planned and facilitated > 250 lay advisory board meetings. A community-based ethics training, similar to what was used in our prior work will be administered to all PAs prior to starting the training for Peers LEAD.

### Intervention Fidelity

As a means to evaluate intervention fidelity – the extent to which the intervention is delivered as it was intended[55] – we will utilize the following process: (1) audio recording group sessions to determine how the intervention is implemented and (2) weekly phone calls from study coordinators to PAs and PBs to discuss content of phone calls and frequency of calls.

## Data collection and outcome measures

### Quantitative measures

To provide preliminary data on the program’s impact on the primary outcome, changes in medication adherence, as well as secondary outcomes (illness beliefs, beliefs about diabetes medicines, self-efficacy, diabetes related psychosocial self-efficacy, health literacy, patient activation, we will collect data from the PBs using a 20-minute paper survey at three points: prior to and upon completion of the 8-week intervention and one month following the completion of the intervention. PBs’ A1c and blood pressure will also be measured before and after participation in Peers LEAD as a clinical indicator of changes in medication adherence. A study team researcher with clinical experience will perform a finger prick test to collect a small amount of blood to test for blood sugars. A1c+ now glucometer will be used to measure the participant’s A1c level while Omron Healthcare Inc. 7-Series Upper Arm Blood Pressure Monitor will be used to measure the blood pressure. The research team will promptly report all blood glucose levels and blood pressure readings to the participant by writing the result on a paper. This will be offered to the participant discreetly. Since glucometers are widely available to the public for self-testing, specialized qualifications are not required for the A1c testing and reporting for this study. Clinical protocols for both A1c and blood pressure will be developed by the study team and approved by a physician study team member. Table 2 provides details on the measures. Demographic data that will be collected include age, gender, race, as well as clinical factors: self-reported health, number of medications used will be collected.

**Table 2.** Quantitative Outcome Measures used in the Peers LEAD intervention.

| **Feasibility outcomes** | **Measures** |
| --- | --- |
| PA/PB recruitment | Recruitment statistics |
| PA/PB attrition | Initial/Final meeting attendance records |
| PA/PB participation in training and intervention sessions | Meeting/training attendance records and feedback from participants |
| **Intervention Outcomes** | **Measures** |
| *Primary* |  |
| Blood glucose | HbA1c |
| Medication adherence | ARMS-D Scale[53] |
| *Secondary* |  |
| Blood pressure | Systolic and Diastolic blood pressure |
| Diabetes health beliefs | Brief Illness Perception Questionnaire (BIPQ)[56] |
| Beliefs about diabetes medicines | Beliefs about Medicines Questionnaire (BMQ) [57] |
| Diabetes medication self-efficacy (SE) | Self-Efficacy for Adherence to Medication Scale (SEAMS) [58] |
| Diabetes psychosocial SE | Diabetes Empowerment Scale – Short Form (DES-SF) [59] |
| Health Literacy | Newest Vital Sign (NVS) [60] |
| Patient provider communication | Patient’s Perceived Involvement in Care Scale[61] |
| Social Support | Social Support section of Diabetes Care Profile [62] |

To examine for larger scalability, several feasibility outcomes will be evaluated: successful recruiting of PAs and PBs, PA and PB attrition, and participation in program components (Table 2). We will record the attendance of PAs and PBs at each meeting and intervention session. The number of PAs and PBs who maintained their participation will be compared with the PAs and PBs recruited at the onset.

### Qualitative interviews

Semi-structured interviews will be conducted with all PBs upon completion of the Peers LEAD to get their feedback on the intervention, its impact on changes in adherence and secondary outcomes. A minimum sample of 10 is appropriate for qualitative interviews.[63, 64] Qualitative methods provide rich and detailed information about individuals experience of events.[22] Phenomenology, appropriate in describing an activity, will be used for the interviews.[65] Interviews allow us to clarify questions with participants and provide detail about their view of the intervention and process.[64] The interview characteristics are described in Table 3. Sample questions are provided in Table 4.

**Table 3.** Semi-structured Interview Characteristics.

|  |  |
| --- | --- |
| **Who?** | One-on-one face-to-face with 30 participants. |
| **Why?** | Discuss the intervention processes, outcomes, and intervention experience. |
| **What?** | What are their views of the intervention, its impact, the intervention's feasibility, acceptability, delivery, and content? |
| **How long/where?** | 60-minute, private locations in two cities |
| **How?** | Audio recorded sessions with Principal Investigator/ Research specialist. |

**Table 4.** Sample Interview Questions.

|  |
| --- |
| How easy/hard was finding a time to talk with your peer ambassador? |
| Did you feel comfortable talking to your peer ambassador? |
| Was it helpful talking to a peer over the phone? |
| Are there different ways you would have liked the calls structured? |
| If you could change Peers LEAD, what would you change? |
| What part of Peers LEAD was of most/least benefit to you? |

### Focus Groups

All PAs at each city will be invited to participate in a 90-minute focus group upon completion of the 8-week intervention. Compared to individual interviews, focus groups allow a range and depth of responses and PAs can stimulate new thoughts for each other, which might otherwise not occur. Questions will elicit feedback about feasibility outcomes: their experiences with the recruitment process, training session, maintaining participation during Peers LEAD, and what would make working as a PA easier.

## Data analysis plan

Since this is a pilot study, there was no power analysis or sample size determination. The following outlines our data analysis plan for both quantitative and qualitative data.

### Quantitative

Paired t-tests will examine pretest vs. post-test changes in PB’s self-reported medication adherence, A1c and other secondary outcomes. Descriptive statistics will be calculated for three feasibility measures related to PAs/PBs: recruitment of PAs, attrition for PAs and extent of PA participation in training and intervention sessions. The recruitment approach will be considered feasible if: there is recruitment of all PAs and PBs as planned, attrition for PAs/PBs is 10% or less, and the participation rate for PAs and PBs is 80% or higher.

### Qualitative

Qualitative content analysis with NVivo 10 will be used to organize themes. The transcripts will be initially read to achieve immersion; data will be read line by line; codes will be created; themes will be developed and organized; and a conceptual model for how the themes are linked will be developed. [66] A comparison of themes will explore the similarities, differences, and interconnections. Analysis will occur until data saturation, i.e., we cannot find new dimensions within the data.[65, 67, 68] A research assistant and PI will code the transcripts independently, discuss similarities and divergences and reach agreement. To check for accuracy, a result summary will be given to 4 participants (based on indicated interest) to conduct member checking.[69]

## Discussion

To our knowledge, this is the first study to develop and implement a culturally appropriate peer-supported educational-behavioral medication adherence intervention for AAs with type 2 diabetes. We use a culturally tailored combination of interdisciplinary education and peer support to address the specific behavioral (e.g., self-efficacy and culturally specific beliefs about diabetes), social (e.g. peer support), and structural factors (e.g., lack of access to information, mistrust of providers) identified from our prior work with AAs with type 2 diabetes to influence medication adherence.[39, 70] This research will advance the understanding of the mechanisms operating within medication management for AAs. This study utilizes a community-engaged collaborative approach involving patient stakeholders throughout the research process by directly engaging AAs with diabetes who are successfully taking their medicines; to utilize their experience, knowledge and advice.[71] In the initial planning phase of Peers LEAD, our PAB (advisory board) provided feedback by informing and refining intervention components, as well as serving as peer mentors. They will also be engaged in the dissemination of results. This study methodologically advances the design of interventions for AAs by initially utilizing theories based on minority health and health disparities science to explore perceptions of diabetes and medications[39], which identified constructs to focus on in the intervention. We utilized similar theories to inform the intervention toolkit and are continuing in the intervention implementation using the same theoretical frameworks.

This study builds upon our prior studies of AAs with type 2 diabetes’ experiences with diabetes and medication adherence.[24, 39, 70] We showed that there were diabetes and medication misperceptions resulting from a lack of knowledge and/or confusion. AAs felt a loss of autonomy because of diabetes. Reasons for nonadherence were fear and frustration due to taking medicines and a disbelief of diabetes diagnosis. Perceived solutions for improving adherence included counseling on the necessity of medicines, and the need for an AA buddy system to support and teach self-advocacy to enhance self-confidence in self-management.[24] Perceived coping behaviors that could enhance AAs’ control of diabetes included empowerment and positive attitudes, and support from peers.[39] In our prior study, some AAs did not take their medicines because of the unavailability of resources to help with self-management, and a lack of knowledge on how to ask their provider questions. [24] Navigating the health care system to access available resources also seemed difficult. By enhancing AAs ability for navigation, thus improving health literacy, we expect an increase in AAs self-efficacy and activation; patient engagement and motivation to take active roles in their self-management and shared decision making with providers.

To develop Peers LEAD, we used an intervention mapping framework for needs assessment (using literature review and six focus groups), drafted the 8-week theory-based program strategies and toolkit with review from our patient advisory board, and an evaluation plan to examine the intervention effects. As a first phase in Peers LEAD, we carefully designed and tested the first 3 weeks to evaluate its feasibility.[54] This was essential before developing further content for the remaining 5 weeks. Results showed PBs enjoyed their shared diabetes experiences with their PA, discussing medication management strategies and navigating the healthcare system. All PB’s liked the easy availability of the PA and reported a positive, trusting relationship between them. [54]

The research team anticipates that there will be challenges throughout the study and has strategized on how to address these challenges, when possible. Recruiting enough AAs may prove to be challenging. To address this, the study team has developed strong partnerships with several community-based organizations in each of the study locations. Early in the process of designing the study, the study team met with each organization to present the proposed project and obtained their commitment to assist with recruiting. Another potential challenge is retaining program participants throughout the intervention. To ensure maximum participation and retention of participants, the study team will be implementing weekly check-in phone calls with each PA and PB. These calls will serve two purposes: (1) to assess for intervention fidelity and(2) to offer support to the PAs and PBs in navigating their way through the phone calls. The intention is to enhance and promote continued participation by having the study team have frequent contact with the PAs and PBs.

By utilizing patient feedback to refine Peers LEAD and piloting it to examine its feasibility, we will generate evidence regarding its real world use and provide support for a randomized controlled trial of its impact on AAs diabetes medication adherence and clinical outcomes. As well, these data will support a future large-scale randomized controlled trial to test the effectiveness of embedding Peers LEAD into a diabetes self-management program for AAs.

## Data Availability

No data is available since this is a protocol paper

## Acknowledgements

We would like to thank the community advisory board members who endorsed this study. This project is supported by the Clinical and Translational Science Award (CTSA) program, through the National Institutes of Health (NIH) National Center for Advancing Translational Sciences, grant UL1TR002373–02. The content is solely the responsibility of the authors and does not necessarily represent the official views of the NIH.

